# Associations between different intensities of leisure-time physical activity and subsequent psychotropic medication use among young and early midlife adults: a register-linked follow-up study

**DOI:** 10.64898/2026.11.11.26367258

**Authors:** Lari Järvensivu, Ville Päivärinne, Ossi Rahkonen, Tea Lallukka

## Abstract

**Introduction:** Previous studies have not examined associations between physical activity intensity and subsequent psychotropic medication use. We examined associations between different leisure-time physical activity (LTPA) intensities and subsequent psychotropic medication purchases.

**Methods:** We used Helsinki Health Study questionnaire data collected in 2017 from 19–39-year-old employees of the City of Helsinki, linked to register data on psychotropic medication purchases from 2014 to 2022 (82.5% consented to the register linkage). The analytical sample included 4,160 participants (80.4% women, representing the target population). Weekly hours of light (LPA), moderate (MPA) and vigorous (VPA) LTPA were examined separately. Cox proportional hazards regression with restricted cubic splines was used to examine nonlinear associations with the first psychotropic medication purchase during follow-up, adjusting for prior medication and sociodemographic and health-related covariates. Hazard ratios (HRs) and 95% confidence intervals (CIs) were reported.

**Results:** During the follow-up, 28.1% of participants had at least one psychotropic medication purchase. After adjustment for age and gender, VPA was associated with psychotropic medication purchases (overall p<0.001; nonlinearity p<0.001), whereas MPA and LPA were not. Further adjustments and stratification by prior psychotropic medication purchase status attenuated the VPA association (overall *p*=0.0041). In this model, for 5 versus 0 hours/week of VPA, the HR was 0.79 (95% CI 0.68–0.91), corresponding to approximately 23 fewer first-purchase events per 1,000 participants over three years.

**Conclusions:** Higher-intensity LTPA appeared more strongly associated with a lower hazard of psychotropic medication purchases, although only the VPA association saw clear support.

## Introduction

According to the Global Burden of Disease study (GBD), mental disorders are among the top ten causes of disease burden (Santomauro et al., 2026). In Finland, among young and working-age adults, aged 20–39, psychological distress has increased from 24.6% in 2018 to 33.7% in 2024 (Parikka et al., 2025). To treat mental disorders, psychotropic medications are increasingly prescribed. Globally, psychotropic medications have seen an average annual increase of 4.1% in sales between 2008 and 2019 (Brauer et al., 2021). Similarly, in Finland, the prevalence of at least one annual psychotropic medication purchase has risen from 7.7% to 16.3% between 2014 and 2025 among 18–29-year-olds (Kela, n.d.). In addition to individual suffering, psychotropic medications reflect the burden on health care associated with mental disorders and can induce hazardous side effects, such as weight gain and dyslipidemia (Anthamatten & Henry-Okafor, 2025). Together, these factors highlight the need for practical and effective interventions that could lessen the need for psychotropic medications.

Results from both longitudinal observational and experimental studies suggest that physical activity is beneficial for mental health (Munro et al., 2026; Rahmati et al., 2024). Additionally, a recent Cochrane review could not establish a clear difference in effectiveness between physical exercise and antidepressant medication in the treatment of depression (Clegg et al., 2026). However, less is known about how different intensities of physical activity are associated with subsequent psychotropic medication use.

Wolf et al. (2024) conducted a meta-analysis investigating the association between physical activity and subsequent psychotropic medication use. The pooled estimates from their main analysis of three studies (N = 40,111) indicated that engagement in any physical activity was associated with a 15% lower likelihood of subsequent psychotropic medication use compared to inactivity (Donoghue et al., 2018; Havnen et al., 2023; Sheikh et al., 2018). Their secondary analysis of two studies found that higher vs. lower physical activity (using cut-offs of 5 hours/week and 14 MET-hours/week in the two respective studies) was associated with a 28% lower risk of future psychotropic medication use (Kleppang et al., 2019; Lahti et al., 2013). These findings provide limited insight into how physical activity relates to subsequent psychotropic medication use, as binary categorization of total activity may mask differences between intensity levels and fail to capture nonlinear dose–response associations. This is particularly relevant given evidence that different intensities of both self-reported and objectively measured physical activity show nonlinear associations with the incidence of common mental disorders (Ho et al., 2022; Wang et al., 2025). Furthermore, a recent study indicated nonlinear dose–response relationships between total LTPA and the use of several different classes of psychotropic medication, which were examined separately (Hernandez et al., 2025).

Examining physical activity intensities as continuous exposures and allowing nonlinear associations can provide a more detailed understanding of the relationship between physical activity and subsequent psychotropic medication use. Using the Helsinki Health Study (HHS) data from young and early midlife employees, the present study aims to address this gap by examining light-, moderate-, and vigorous-intensity leisure-time physical activity (LTPA) as separate, mutually adjusted continuous variables and using nonlinear modeling to investigate their associations with subsequent prescribed, reimbursed psychotropic medication purchases. As prior psychotropic medication purchases and other important covariates may affect the associations between leisure-time physical activity and psychotropic medication use, these factors were considered in the analyses (Choi et al., 2017; Hernandez et al., 2025; Korslund et al., 2023; Wolf et al., 2024).

Specifically, we aimed to answer the following research question:

> How are different intensities of LTPA associated with a first psychotropic medication purchase during follow-up in young and early midlife adults, controlling for key covariates, and what are the shapes of these associations?

## Methods

### Sample

We used HHS survey data collected from employees of the City of Helsinki, Finland. The survey includes a large array of questions relating to general health and work ability. In 2017, the survey was sent to all 19–39-year-olds who had been employed for at least 4 months and were working half-time or more for the City of Helsinki (*n*=11,459). Responding to the survey was possible either online through a secure server or using a mailed survey (Lallukka et al., 2020). A shorter telephone interview was conducted for non-respondents, resulting in a final response rate of 51.5% (*n*=5898, 78.5% women). For the present study, the telephone respondents (*n*=787) were excluded because the shorter interview did not include questions on LTPA.

These survey data were linked with the Social Insurance Institution of Finland (Kela) register data from 2014 to 2022 on psychotropic medication purchases (82.5% gave consent for register linkages). After excluding those without LTPA data, the final analytical sample consisted of 4,160 participants (80.4% women). This process is depicted in further detail in Figure 1.

**Figure 1.**
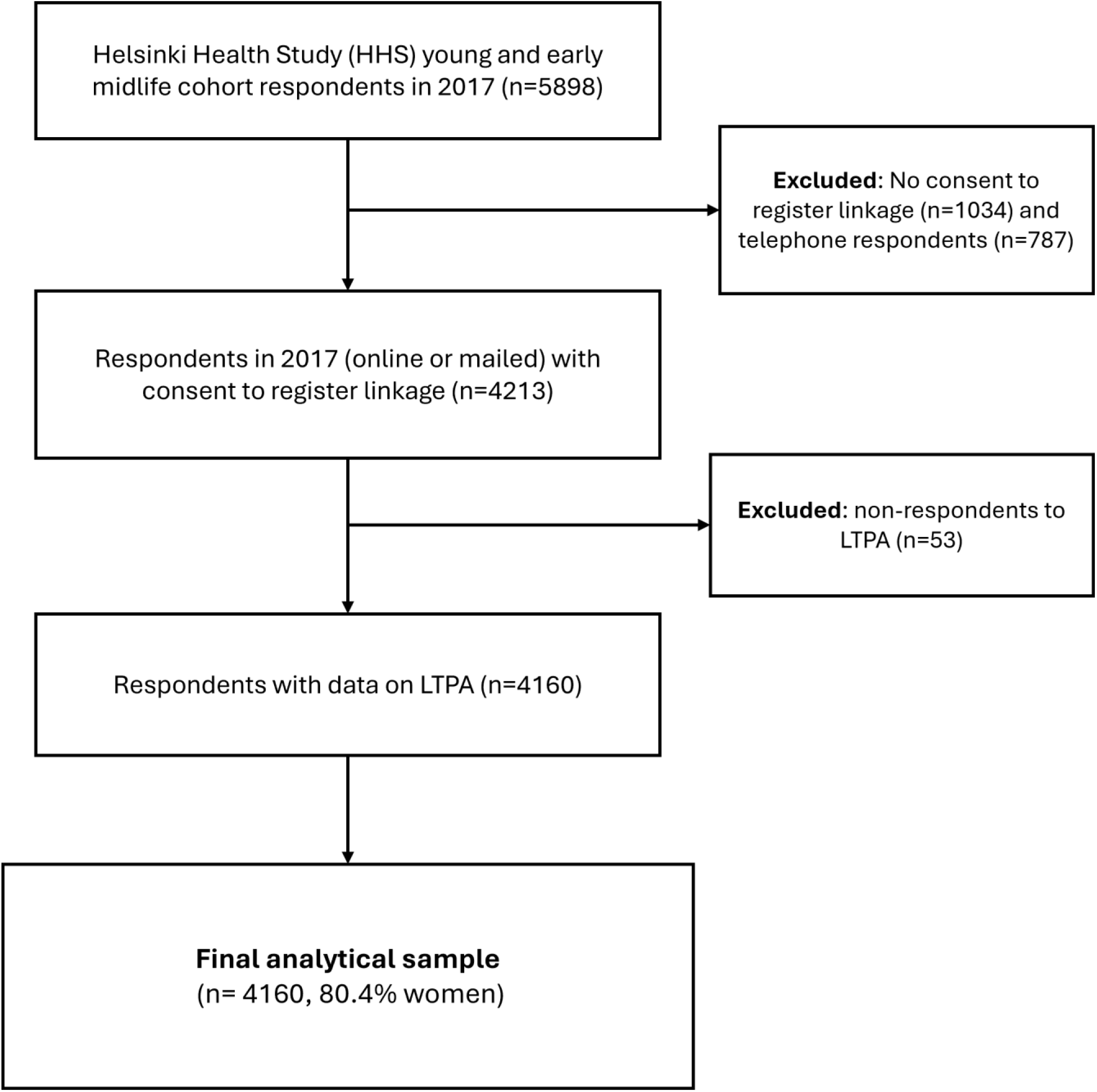
Flow chart of participant exclusions leading to the final analytical sample (n=4,160) among 19–39-year-old participants of the Helsinki Health Study *Note. L*TPA = leisure-time physical activity.

### Leisure-time physical activity

In the 2017 survey, participants were asked to report the weekly time spent at four different intensity levels: (1) walking or equivalent activities, (2) brisk walking or equivalent activities, (3) light running (jogging) or equivalent activities, and (4) brisk running or equivalent activities. The durations were reported separately for commuting activity and other LTPA but were subsequently combined because the two domains showed very similar associations when analyzed separately (Figure S1). Three intensity-specific LTPA variables were formed: weekly hours of light physical activity (LPA), corresponding to walking or activities of equivalent intensity; weekly hours of moderate physical activity (MPA), corresponding to brisk walking or activities of equivalent intensity; and weekly hours of vigorous physical activity (VPA), combining running/jogging, running, and other activities of equivalent intensity.

For descriptive purposes, metabolic equivalent (MET) hours were calculated for LPA, MPA, and VPA by multiplying the hours spent at each intensity by their corresponding MET values. The exact MET multipliers and calculations are detailed in Table S1. MET-hours of moderate-to-vigorous physical activity (MVPA), combining MPA and VPA, and total LTPA, combining all intensity levels, were also calculated for descriptive purposes and sensitivity analyses. LPA, MPA, and VPA were additionally categorized for descriptive purposes.

### Psychotropic medication

We used register information on prescribed reimbursed psychotropic medication purchases from the Social Insurance Institution of Finland. Data from this register are classified according to the WHO Anatomical Therapeutic Chemical (ATC) Classification System (WHO, n.d.). We included psychotropic medications coded as N05 (psycholeptics) and N06 (psychoanaleptics), excluding N06D, which comprises antidementia drugs.

For participants who had given informed written consent (n=4213), the information on psychotropic purchases was linked to the 2017 survey data. We had information on prior psychotropic medication purchases for the three years preceding the survey. Participants were followed from the date of survey response until their first recorded psychotropic medication purchase or the end of 2022, whichever occurred first. For descriptive purposes, participants were additionally grouped according to the number of psychotropic medication purchases during follow-up (0, 1–2, and ≥3 purchases).

### Covariates

The covariates included potentially relevant sociodemographic, work- and health-related factors. These were chosen based on their previously reported associations with LTPA and mental health and/or psychotropic medication (Choi et al., 2017; Hernandez et al., 2025; Korslund et al., 2023; Wolf et al., 2024). We included information on participants’ gender, age, education, marital status, working conditions, alcohol and tobacco consumption, fresh vegetable consumption, body mass index (BMI) and sedentary time, all derived from the 2017 survey responses. Education level comprised a three-category variable: ‘low’ (upper secondary school or lower), ‘intermediate’ (bachelor’s degree) and ‘high’ (master’s degree or higher). Marital status was dichotomized into ‘married or cohabiting’ and ‘not married or cohabiting’ based on five response alternatives. Physical and mental strenuousness of work were both dichotomized into ‘high’ (very strenuous or rather strenuous) and ‘low’ (rather light or very light) groups, based on four response alternatives. Binge drinking was defined as consuming more than six portions of alcohol on one occasion at least once a week for men and once a month for women, dividing participants into ‘binge drinkers’ and ‘not binge drinkers’. Tobacco consumption was divided into ‘daily or occasional’ and ‘non- or ex-smokers’. Fresh vegetable consumption was dichotomized as ‘daily’ and ‘not daily’. BMI was calculated from participants’ self-reported height and weight and missing values (*n*=38, 0.9%) were replaced with the median BMI of the analytical sample. Complete information for participants’ age and juridical gender were acquired from the register using the personal identification numbers assigned to each citizen (at birth) and thus there were no missing data on these variables. There were no missing data for sedentary time. Among the remaining covariates, the number of missing observations ranged from 8 (0.19%) for education to 153 (3.68%) for binge drinking. Missing values for categorical covariates were assigned to the category expected to have the lowest risk, following previous procedures (Etholén et al., 2022).

### Statistical analyses

Baseline characteristics were described using means and standard deviations (SDs) for continuous variables and counts and percentages for categorical variables. Differences in baseline characteristics were examined using one-way analysis of variance (ANOVA) for continuous variables and Pearson’s chi-square tests for categorical variables. Categorized LTPA variables were used for descriptive purposes (Table 1).

**Table 1.**
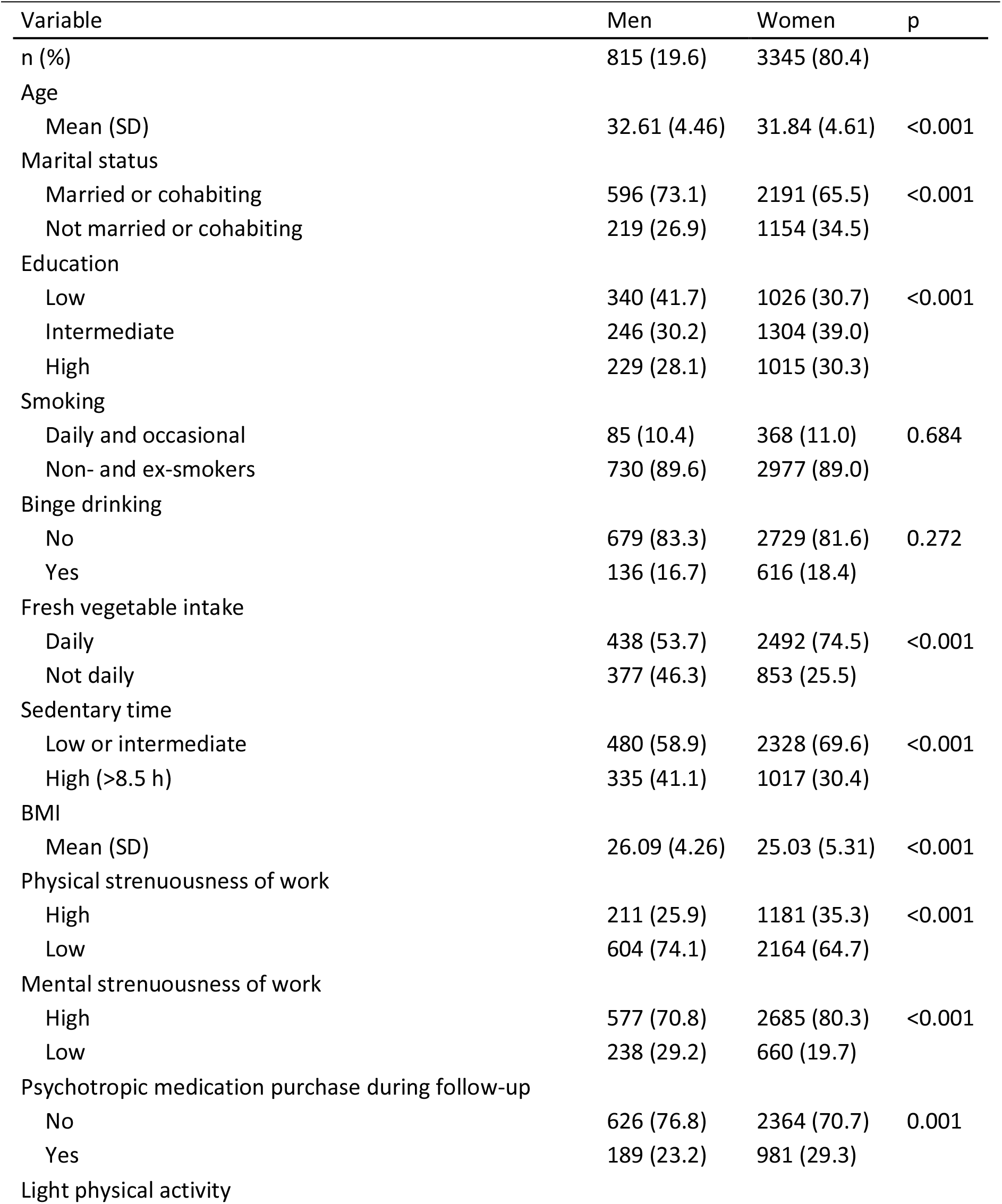
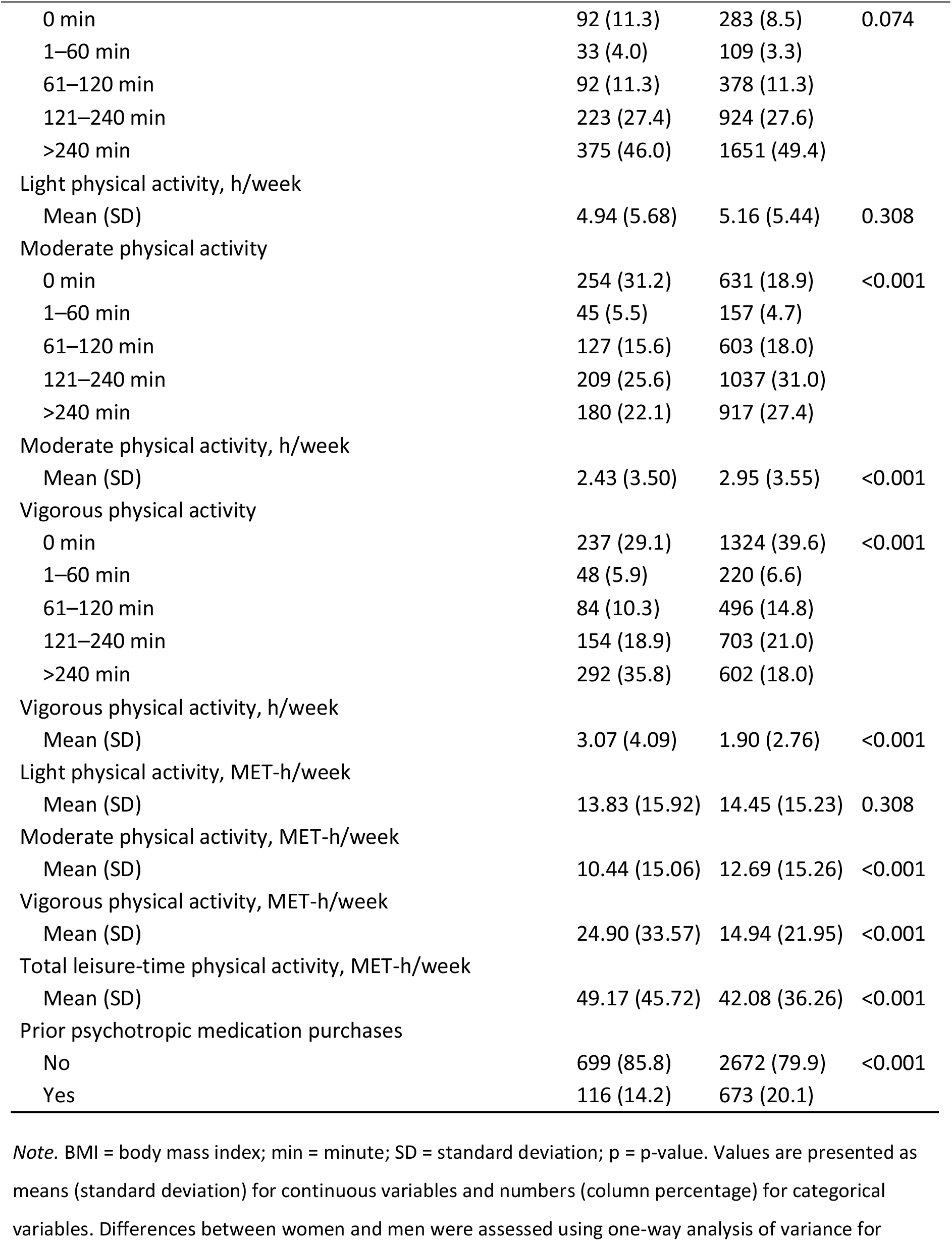
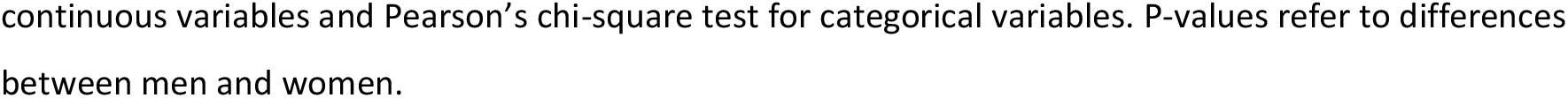
Baseline characteristics in 2017 and psychotropic medication purchases before and during follow-up in 2014–2022, stratified by gender among 19–39-year-old participants of the Helsinki Health Study (N = 4,160)

| Variable | Men | Women | p |
|---|---|---|---|
| n (%) | 815 (19.6) | 3345 (80.4) |  |
| Age |  |  |  |
| Mean (SD) | 32.61 (4.46) | 31.84 (4.61) | <0.001 |
| Marital status |  |  |  |
| Married or cohabiting | 596 (73.1) | 2191 (65.5) | <0.001 |
| Not married or cohabiting | 219 (26.9) | 1154 (34.5) |  |
| Education |  |  |  |
| Low | 340 (41.7) | 1026 (30.7) | <0.001 |
| Intermediate | 246 (30.2) | 1304 (39.0) |  |
| High | 229 (28.1) | 1015 (30.3) |  |
| Smoking |  |  |  |
| Daily and occasional | 85 (10.4) | 368 (11.0) | 0.684 |
| Non- and ex-smokers | 730 (89.6) | 2977 (89.0) |  |
| Binge drinking |  |  |  |
| No | 679 (83.3) | 2729 (81.6) | 0.272 |
| Yes | 136 (16.7) | 616 (18.4) |  |
| Fresh vegetable intake |  |  |  |
| Daily | 438 (53.7) | 2492 (74.5) | <0.001 |
| Not daily | 377 (46.3) | 853 (25.5) |  |
| Sedentary time |  |  |  |
| Low or intermediate | 480 (58.9) | 2328 (69.6) | <0.001 |
| High (>8.5 h) | 335 (41.1) | 1017 (30.4) |  |
| BMI |  |  |  |
| Mean (SD) | 26.09 (4.26) | 25.03 (5.31) | <0.001 |
| Physical strenuousness of work |  |  |  |
| High | 211 (25.9) | 1181 (35.3) | <0.001 |
| Low | 604 (74.1) | 2164 (64.7) |  |
| Mental strenuousness of work |  |  |  |
| High | 577 (70.8) | 2685 (80.3) | <0.001 |
| Low | 238 (29.2) | 660 (19.7) |  |
| Psychotropic medication purchase during follow-up |  |  |  |
| No | 626 (76.8) | 2364 (70.7) | 0.001 |
| Yes | 189 (23.2) | 981 (29.3) |  |
| Light physical activity |  |  |  |
| 0 min | 92 (11.3) | 283 (8.5) | 0.074 |
| 1–60 min | 33 (4.0) | 109 (3.3) |  |
| 61–120 min | 92 (11.3) | 378 (11.3) |  |
| 121–240 min | 223 (27.4) | 924 (27.6) |  |
| >240 min | 375 (46.0) | 1651 (49.4) |  |
| Light physical activity, h/week |  |  |  |
| Mean (SD) | 4.94 (5.68) | 5.16 (5.44) | 0.308 |
| Moderate physical activity |  |  |  |
| 0 min | 254 (31.2) | 631 (18.9) | <0.001 |
| 1–60 min | 45 (5.5) | 157 (4.7) |  |
| 61–120 min | 127 (15.6) | 603 (18.0) |  |
| 121–240 min | 209 (25.6) | 1037 (31.0) |  |
| >240 min | 180 (22.1) | 917 (27.4) |  |
| Moderate physical activity, h/week |  |  |  |
| Mean (SD) | 2.43 (3.50) | 2.95 (3.55) | <0.001 |
| Vigorous physical activity |  |  |  |
| 0 min | 237 (29.1) | 1324 (39.6) | <0.001 |
| 1–60 min | 48 (5.9) | 220 (6.6) |  |
| 61–120 min | 84 (10.3) | 496 (14.8) |  |
| 121–240 min | 154 (18.9) | 703 (21.0) |  |
| >240 min | 292 (35.8) | 602 (18.0) |  |
| Vigorous physical activity, h/week |  |  |  |
| Mean (SD) | 3.07 (4.09) | 1.90 (2.76) | <0.001 |
| Light physical activity, MET-h/week |  |  |  |
| Mean (SD) | 13.83 (15.92) | 14.45 (15.23) | 0.308 |
| Moderate physical activity, MET-h/week |  |  |  |
| Mean (SD) | 10.44 (15.06) | 12.69 (15.26) | <0.001 |
| Vigorous physical activity, MET-h/week |  |  |  |
| Mean (SD) | 24.90 (33.57) | 14.94 (21.95) | <0.001 |
| Total leisure-time physical activity, MET-h/week |  |  |  |
| Mean (SD) | 49.17 (45.72) | 42.08 (36.26) | <0.001 |
| Prior psychotropic medication purchases |  |  |  |
| No | 699 (85.8) | 2672 (79.9) | <0.001 |
| Yes | 116 (14.2) | 673 (20.1) |  |

*Note.* BMI = body mass index; min = minute; SD = standard deviation; p = p-value. Values are presented as means (standard deviation) for continuous variables and numbers (column percentage) for categorical variables. Differences between women and men were assessed using one-way analysis of variance for continuous variables and Pearson’s chi-square test for categorical variables. P-values refer to differences between men and women.

The main analyses used Cox proportional hazards regression with restricted cubic splines (RCSs) (Austin, 2025; Govindarajulu et al., 2009). LPA, MPA and VPA were included simultaneously as predictors, allowing for mutual adjustment, and time to the first psychotropic medication purchase was used as the outcome variable. Follow-up time was calculated from the return date of the 2017 survey until the first registered psychotropic medication purchase or end of 2022, whichever occurred first. Participants without a registered psychotropic medication purchase during follow-up were treated as censored at the end of follow-up. RCS functions were incorporated to examine nonlinear dose–response associations. In all analyses, three knots were used, with knots placed at the 10^th^, 50^th^, and 90^th^ percentiles following Harrell’s (2015) default knot-placement procedure.

Multicollinearity and the proportional hazards assumption were assessed. LPA, MPA, and VPA were weakly to moderately correlated, with the strongest correlation between LPA and MPA (*r* = 0.40) (Table S2). The degrees-of-freedom-adjusted generalized variance inflation factors (GVIF^1/(2*df*)^) for LPA, MPA and VPA were low (approximately 1.9–2.1), while the values for all covariates were below 1.32 (Table S3). Overall, there was no evidence of problematic multicollinearity. The proportional hazards assumption was assessed using the Grambsch–Therneau test (1994) (Table S4) and scaled Schoenfeld residuals. The Grambsch–Therneau test indicated potential nonproportional hazards for several covariates, particularly prior psychotropic medication purchase status (p < 0.001), and for LPA (p = 0.0267). However, given the sensitivity of formal tests of proportional hazards to small departures in large samples, graphical diagnostics were considered alongside the statistical tests.

The Schoenfeld residual plots did not indicate a clear or meaningful violation of the proportional hazards assumption for LPA or the covariates, apart from prior psychotropic medication purchase status, for which the estimated coefficient clearly decreased over the follow-up period (Figure S2). Consequently, because prior psychotropic medication purchase status was not an exposure of interest and no effect estimate was required for this covariate, the models were stratified by prior psychotropic medication purchase status. Model 1 was adjusted for gender and age. The baseline hazard function was additionally stratified by prior psychotropic medication purchase status in Model 2. Sociodemographic, health- and work-related factors were further adjusted for in Model 3, while keeping the baseline hazard function stratified by prior psychotropic medication purchase status.

The primary analyses were conducted in the pooled sample with adjustment for gender, while analyses including gender interaction terms were performed as sensitivity analyses to assess the robustness and potential heterogeneity of the associations (Table S9). This was because the relatively small number of men (n = 815) in our sample made the male-specific estimates less precise.

All analyses were conducted using R version 4.4.1 (R Core Team, 2024). For the main analyses, hazard ratios (HRs) and corresponding 95% confidence intervals (CIs) were calculated. Additionally, model-derived standardized three-year risks of a first psychotropic medication purchase were estimated by averaging participant-specific predicted risks from the Cox models while retaining observed values for all other covariates. Absolute risk differences were expressed per 1,000 participants over three years. Two-sided tests were used, and statistical significance was set at *p* < 0.05.

## Results

The study included 4,160 participants (80.4% women) with a mean age of 32.0 years (SD = 4.59). Men were slightly older and had a higher mean BMI than women. Men were also more likely to have lower educational attainment, be married or cohabiting, not daily fresh vegetable consumption, and report high sedentary time. Conversely, men were less likely to report physical or mental strenuousness of work. No evidence of gender differences was observed in smoking, binge drinking, or LPA. Women reported more MPA, whereas men reported more VPA. Women were more likely to have prior psychotropic medication purchases and to purchase psychotropic medication during follow-up. Further characteristics of the sample are presented in Table 1.

Table 2 shows differences between participants who purchased and did not purchase psychotropic medication during the 2017–2022 follow-up. During follow-up, 1,170 participants (28.1%) had at least one registered psychotropic medication purchase.

**Table 2.** Baseline characteristics in 2017 by number of psychotropic medication purchases during follow-up among 19–39-year-old participants of the Helsinki Health Study (N = 4,160)

| Variable | No purchases | 1–2 purchases | 3 or more purchases | p |
|---|---|---|---|---|
| n (%) | 2990 (71.9) | 336 (8.1) | 834 (20.0) |  |
| Gender |  |  |  |  |
| Women | 2364 (79.1) | 284 (84.5) | 697 (83.6) | 0.002 |
| Men | 626 (20.9) | 52 (15.5) | 137 (16.4) |  |
| Age |  |  |  |  |
| Mean (SD) | 31.98 (4.59) | 31.98 (4.68) | 32.03 (4.56) | 0.948 |
| Marital status |  |  |  |  |
| Married or cohabiting | 2101 (70.3) | 220 (65.5) | 466 (55.9) | <0.001 |
| Not married or cohabiting | 889 (29.7) | 116 (34.5) | 368 (44.1) |  |
| Education |  |  |  |  |
| Low | 909 (30.4) | 128 (38.1) | 329 (39.4) | <0.001 |
| Intermediate | 1139 (38.1) | 106 (31.5) | 305 (36.6) |  |
| High | 942 (31.5) | 102 (30.4) | 200 (24.0) |  |
| Physical strenuousness of work |  |  |  |  |
| High | 972 (32.5) | 117 (34.8) | 303 (36.3) | 0.101 |
| Low | 2018 (67.5) | 219 (65.2) | 531 (63.7) |  |
| Mental strenuousness of work |  |  |  |  |
| High | 2311 (77.3) | 270 (80.4) | 681 (81.7) | 0.017 |
| Low | 679 (22.7) | 66 (19.6) | 153 (18.3) |  |
| Smoking |  |  |  |  |
| Daily or occasional smoker | 269 (9.0) | 40 (11.9) | 144 (17.3) | <0.001 |
| Non- or ex-smoker | 2721 (91.0) | 296 (88.1) | 690 (82.7) |  |
| Binge drinking |  |  |  |  |
| No | 2514 (84.1) | 266 (79.2) | 628 (75.3) | <0.001 |
| Yes | 476 (15.9) | 70 (20.8) | 206 (24.7) |  |
| Fresh vegetable intake |  |  |  |  |
| Daily | 2153 (72.0) | 233 (69.3) | 544 (65.2) | 0.001 |
| Not daily | 837 (28.0) | 103 (30.7) | 290 (34.8) |  |
| Sedentary time |  |  |  |  |
| Low or intermediate | 2050 (68.6) | 228 (67.9) | 530 (63.5) | 0.024 |
| High (>8.5 h/day) | 940 (31.4) | 108 (32.1) | 304 (36.5) |  |
| BMI |  |  |  |  |
| Mean (SD) | 24.99 (4.90) | 24.95 (4.76) | 26.27 (5.92) | <0.001 |
| Light physical activity, h/week |  |  |  |  |
| Mean (SD) | 5.05 (5.47) | 5.51 (5.52) | 5.21 (5.54) | 0.289 |
| Light physical activity category |  |  |  |  |
| 0 min/week | 289 (9.7) | 24 (7.1) | 62 (7.4) | 0.183 |
| 1–60 min/week | 92 (3.1) | 17 (5.1) | 33 (4.0) |  |
| 61–120 min/week | 343 (11.5) | 38 (11.3) | 89 (10.7) |  |
| 121–240 min/week | 827 (27.7) | 85 (25.3) | 235 (28.2) |  |
| >240 min/week | 1439 (48.1) | 172 (51.2) | 415 (49.8) |  |
| Moderate physical activity, h/week |  |  |  |  |
| Mean (SD) | 2.87 (3.44) | 2.81 (3.17) | 2.79 (4.04) | 0.823 |
| Moderate physical activity category |  |  |  |  |
| 0 min/week | 614 (20.5) | 75 (22.3) | 196 (23.5) | 0.332 |
| 1–60 min/week | 137 (4.6) | 15 (4.5) | 50 (6.0) |  |
| 61–120 min/week | 539 (18.0) | 53 (15.8) | 138 (16.5) |  |
| 121–240 min/week | 894 (29.9) | 107 (31.8) | 245 (29.4) |  |
| >240 min/week | 806 (27.0) | 86 (25.6) | 205 (24.6) |  |
| Vigorous physical activity, h/week |  |  |  |  |
| Mean (SD) | 2.27 (3.23) | 1.98 (2.56) | 1.69 (2.75) | <0.001 |
| Vigorous physical activity category |  |  |  |  |
| 0 min/week | 1037 (34.7) | 124 (36.9) | 400 (48.0) | <0.001 |
| 1–60 min/week | 190 (6.4) | 26 (7.7) | 52 (6.2) |  |
| 61–120 min/week | 428 (14.3) | 45 (13.4) | 107 (12.8) |  |
| 121–240 min/week | 645 (21.6) | 73 (21.7) | 139 (16.7) |  |
| >240 min/week | 690 (23.1) | 68 (20.2) | 136 (16.3) |  |
| Light physical activity, MET-h/week |  |  |  |  |
| Mean (SD) | 14.13 (15.31) | 15.43 (15.45) | 14.59 (15.52) | 0.289 |
| Moderate physical activity, MET-h/week |  |  |  |  |
| Mean (SD) | 12.34 (14.78) | 12.07 (13.64) | 11.99 (17.36) | 0.823 |
| Vigorous physical activity, MET-h/week |  |  |  |  |
| Mean (SD) | 18.01 (26.06) | 15.70 (20.51) | 13.39 (22.16) | <0.001 |
| Total leisure-time physical activity, MET-h/week |  |  |  |  |
| Mean (SD) | 44.47 (38.81) | 43.19 (31.35) | 39.97 (39.29) | 0.011 |
| Prior psychotropic medication purchases |  |  |  |  |
| No | 2794 (93.4) | 229 (68.2) | 348 (41.7) | <0.001 |
| Yes | 196 (6.6) | 107 (31.8) | 486 (58.3) |  |

*Note.* MET = metabolic equivalent; min = minute; SD = standard deviation; p = p-value. Values are presented as means (standard deviation) for continuous variables and numbers (column percentage) for categorical variables. Differences between groups were assessed using one-way analysis of variance for continuous variables and Pearson’s chi-square test for categorical variables. p-values refer to overall differences across the three purchase groups.

Participants with one or more psychotropic medication purchases were more likely to be women, have lower education, be unmarried, binge drinkers, and have a history of psychotropic medication purchases than those with no purchases, and these differences appeared generally most pronounced between participants with three or more purchases and participants with no purchases. Those with three or more purchases also had a higher mean BMI and were more likely to smoke, consume fresh vegetables less frequently, report high sedentary time, and experience high mental work strenuousness. LTPA differences were primarily observed for VPA, where participants with 1–2 purchases reported slightly less VPA than those with no purchases, while the lowest levels occurred among those with three or more purchases. Differences in total LTPA were also observed between the purchase groups, whereas LPA and MPA did not differ significantly between the groups.

Restricted cubic spline analyses demonstrated a nonlinear association between VPA and the hazard of first psychotropic medication purchase in the age- and gender-adjusted Model 1 (overall p < 0.0001; p for nonlinearity = 0.0001). The hazard decreased with increasing VPA up to approximately 5 hours/week, at which point the HR was 0.65 (95% CI 0.59–0.71) compared with 0 hours/week, before leveling off (Figure 2). The corresponding model-derived standardized absolute risk difference was approximately 78 fewer participants purchasing psychotropic medication per 1,000 participants over three years.

**Figure 2.**
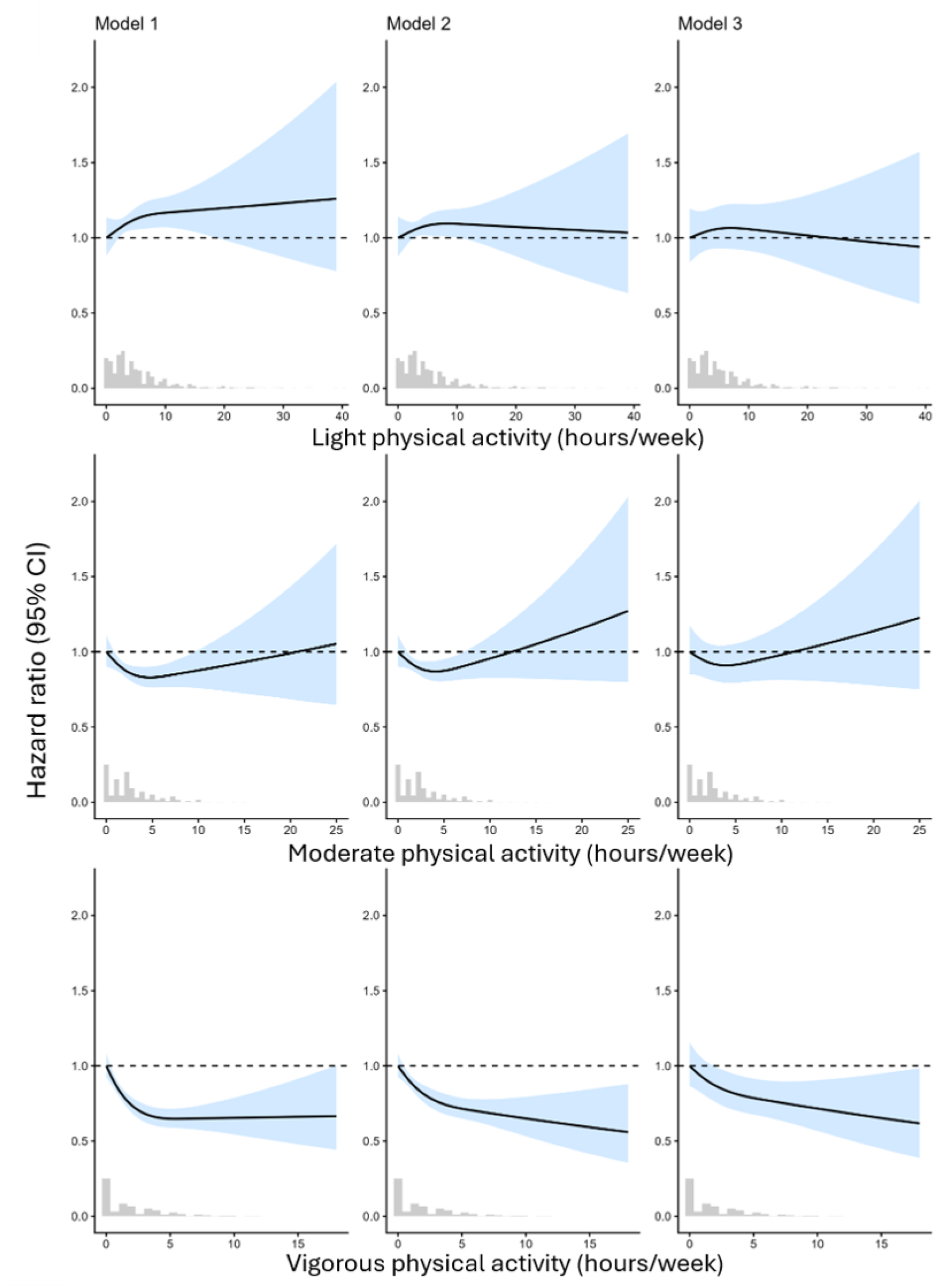
Associations of mutually adjusted light, moderate, and vigorous leisure-time physical activity with the hazard of a first psychotropic medication purchase among 19–39-year-old participants of the Helsinki Health Study (N = 4,160). *Note*. Histograms depict the distribution of participants across values of light, moderate, and vigorous physical activity. Model 1 was adjusted for age and gender. Model 2 was adjusted for age and gender, with the baseline hazard stratified by prior psychotropic medication purchase status. Model 3 was adjusted for age, gender, education, body mass index, smoking, binge drinking, marital status, fresh vegetable intake, sedentary time, physical strenuousness of work, and mental strenuousness of work, with the baseline hazard stratified by prior psychotropic medication purchase status. CI = confidence interval.

After stratification of the baseline hazard by prior psychotropic medication purchase status (Model 2), the association between VPA and the outcome remained statistically significant (overall p < 0.0001; p for nonlinearity = 0.0372) but was slightly attenuated. Further adjustment for sociodemographic, work- and health-related factors (Model 3) attenuated the association, although higher VPA remained associated with a lower hazard of psychotropic medication purchase. At 5 hours/week of VPA, the HR was 0.79 (95% CI 0.68–0.91) compared with 0 hours/week, corresponding to approximately 23 fewer participants purchasing psychotropic medication per 1,000 participants over three years. The overall association remained statistically significant (p = 0.0041), but evidence for nonlinearity was no longer observed (p = 0.2122).

For MPA, there was evidence of nonlinearity in Models 1 and 2 (p = 0.0284 and p = 0.0344, respectively), although the overall associations were not statistically significant. Neither an overall association nor evidence of nonlinearity was observed for MPA in the fully adjusted model (Model 3). No evidence of association was observed for LPA in any of the models (Table 3). The associations between physical activity intensity and the hazard of a first psychotropic medication purchase across all three models are illustrated in Figure 2, with HRs and 95% CIs.

**Table 3.** Overall and nonlinear Wald tests for restricted cubic spline terms of light, moderate and vigorous leisure-time physical activity in mutually adjusted Cox proportional hazards models among 19–39-year-old participants of the Helsinki Health Study (N = 4,160).

| Exposure | Overall Wald χ<sup>2</sup> | df | p | Nonlinear Wald χ<sup>2</sup> | df | p |
|---|---|---|---|---|---|---|
| **Model 1** |  |  |  |  |  |  |
| LPA (hours/week) | 3.06 | 2 | 0.2165 | 0.97 | 1 | 0.3239 |
| MPA (hours/week) | 5.42 | 2 | 0.0666 | 4.8 | 1 | 0.0284 |
| VPA (hours/week) | 34.31 | 2 | <0.0001 | 16.33 | 1 | 0.0001 |
| **Model 2** |  |  |  |  |  |  |
| LPA (hours/week) | 0.88 | 2 | 0.6456 | 0.65 | 1 | 0.4206 |
| MPA (hours/week) | 4.56 | 2 | 0.1022 | 4.48 | 1 | 0.0344 |
| VPA (hours/week) | 22.42 | 2 | <0.0001 | 4.34 | 1 | 0.0372 |
| **Model 3** |  |  |  |  |  |  |
| LPA (hours/week) | 0.52 | 2 | 0.7697 | 0.52 | 1 | 0.4696 |
| MPA (hours/week) | 2.34 | 2 | 0.3106 | 2.22 | 1 | 0.1363 |
| VPA (hours/week) | 10.98 | 2 | 0.0041 | 1.56 | 1 | 0.2122 |

*Note.* χ<sup>2</sup> = Wald chi-square statistic; df = degrees of freedom; p = p-value; LPA = light physical activity; MPA = moderate physical activity; VPA = vigorous physical activity. Overall p values test the overall association of each restricted cubic spline term with the outcome. Nonlinear p values test the nonlinear spline components. Model 1 was adjusted for age and gender. Model 2 was adjusted for age and gender, with the baseline hazard stratified by prior psychotropic medication purchase status. Model 3 was adjusted for age, gender, education, body mass index, smoking, binge drinking, marital status, fresh vegetable intake, sedentary time, physical strenuousness of work, and mental strenuousness of work, with the baseline hazard stratified by prior psychotropic medication purchase status.

### Sensitivity analyses

The results from our sensitivity analyses were generally in line with the findings from our main analyses that showed stronger associations for VPA. To examine the influence of outliers, we conducted analyses in which participants with any values at or above the 99th percentile for LPA, MPA, or VPA were excluded (n = 106). In this analysis, we observed similar but slightly weaker associations for VPA, while no associations could be confirmed for MPA (Table S5).

We also investigated the associations of total LTPA and MVPA with the hazard of a psychotropic medication purchase using MET-hours. The statistical evidence for an overall association was stronger for MVPA and weaker for total LTPA than for VPA (Table S6). Furthermore, the HR curves looked similar for MVPA and VPA, with a slightly larger maximum hazard reduction observed for MVPA (Figure S3). However, after stratifying the baseline hazard function for prior psychotropic medication purchase status and adjusting for other covariates, the statistical evidence indicated a weaker association for MVPA compared to VPA alone (Table S6).

Excluding those with ongoing psychotropic medication (n=334), defined as having purchased psychotropic medication inside the 100 days leading up to the baseline survey, we saw weaker associations, but the overall association for VPA remained statistically significant in the fully adjusted model (Table S7). Similarly, when excluding those with any purchases in the prior 3 years (n=789), there was still significant evidence for VPA even in the fully adjusted model (*p* = .027), and the HR curves looked similar, although less pronounced, to the ones from our main analysis. These results can be seen in further detail in Table S8 and Figure S5.

When adjusting for age, evidence for moderation by gender was observed for the overall association of both LPA and VPA (Table S9). The VPA associations had similar shapes in men and women, but the decline in the estimated hazard was steeper among men (Figure S4). Conversely, when LPA increased the HR decreased among men but increased among women (Figure S4). This gender-specific LPA association was statistically significant only among women (Table S9). Furthermore, after stratifying the baseline hazard by prior psychotropic medication purchase status, no evidence of moderation by gender was observed (Table S9).

## Discussion

In this study, the findings provided the strongest evidence for an inverse association between higher-intensity LTPA, particularly VPA, and subsequent psychotropic medication purchases. VPA showed the clearest inverse association, with an initially nonlinear dose–response pattern. Some evidence of nonlinearity was observed for MPA in the less-adjusted models, but neither MPA nor LPA showed a significant overall association, although the estimates suggested a possible inverse association for MPA. The hazard of a first psychotropic medication purchase decreased with increasing VPA up to approximately 4–5 hours/week, after which the association appeared to plateau. Compared with 0 hours/week, 5 hours/week of VPA corresponded to approximately 78 fewer participants purchasing psychotropic medication per 1000 participants over three years. However, this figure was attenuated to approximately 23 after accounting for prior psychotropic medication purchase status and adjusting for sociodemographic and work- and health-related factors, and there was no more indication of a nonlinear association. These findings were generally supported by sensitivity analyses excluding participants with above 99^th^ percentile values on the LTPA variables or participants who had prior psychotropic medication purchases, as the inverse association for VPA remained evident, although it was attenuated in some analyses (Tables S5, S7, and S8).

Previous studies have thus far mostly examined associations of broad physical activity categories with psychotropic medication, often based on categorizing participants as either inactive or active based on a specific cut-off. These studies have suggested that being active vs. inactive is associated with a lower hazard of psychotropic medication (Wolf et al., 2024). The present study elucidates this by suggesting that higher intensities, especially VPA, may have been responsible for these previous findings. Additionally, the present study adds to the previous literature on different intensities of physical activity and mental health, by examining registered psychotropic medication purchases, which can be viewed as a downstream indicator of clinically recognized mental health problems.

Our results seemed to be broadly in line with a study examining associations of LPA, MPA and VPA with incidence of depression and anxiety and their co-occurrence (Wang et al., 2025). This study used 365,656 and 91,800 participants from the UK Biobank who had data on self-reported physical activity and device-measured physical activity, respectively. The study found disparate associations for self-report vs. objectively measured physical activity. J-shaped associations were found for self-report LPA, MPA and VPA where the risk of common mental disorders reduced up to a point after which the risk started to slightly increase again. However, after full adjustment for covariates none of the self-reported variables was significantly associated with the incidence of common mental disorders. Conversely, Wang et al. (2025) observed generally more L-shaped dose–response associations for the device-measured intensities and clear evidence for these associations was observed across all adjustments. We found broadly similar associations for MPA and VPA. Consistent with our results, Wang et al. (2025) found the strongest evidence of an association for VPA among the self-reported physical activity variables, while also finding some evidence for LPA and MPA in their age- and gender-adjusted models. In contrast to the self-reported variable results from Wang et al. (2025) we found clear evidence of an association for VPA, even when accounting for prior psychotropic medication purchase status and adjusting for sociodemographic, work- and health-related factors. However, it should be noted that Wang et al. (2025) adjusted for histories of multiple common diseases, which could not be adjusted for in the present study.

The most notable difference from the study by Wang et al. (2025) was that we did not find any evidence of an association between LPA and the hazard of psychotropic medication purchases. However, most other previous studies have not found robust associations between LPA and a lower risk of mental ill-health. A previous study found no association between biking/walking, corresponding to LPA in this study, and depression (Soini et al., 2024). In addition, a systematic review of observational studies on LPA and mental ill-health reported varying findings and could not establish a consistent relationship between LPA and mental ill-health (Felez-Nobrega et al., 2021). Additionally, another UK Biobank study using objective measures for physical activity showed no associations of LPA with either anxiety or depression in mutually adjusted models (Ho et al., 2022). Although Wang et al. (2025) used the same UK Biobank data, they analysed a slightly different sample and did not mutually adjust for the activity intensities. These differences may partly explain the contrasting findings for LPA. Considering that LPA, MPA and VPA are correlated with each other, not using mutual adjustment may mean that the estimated association for LPA may partly have reflected its correlation with MPA and VPA in the study by Wang et al. (2025).

Prior studies have also consistently found inverse associations for MPA, in contrast to our findings (Ho et al., 2022; Soini et al., 2024; Wang et al., 2025). In our age- and gender-adjusted sensitivity analysis, MVPA showed the strongest statistical evidence of an overall association. Based on this and the direction of the results for MPA in the main analysis, it could be that all MVPA is associated with a lower hazard of a psychotropic medication purchase, although this association appears to be mainly driven by VPA. However, in line with our main analysis the MVPA association showed greater attenuation than the VPA association, when accounting for prior psychotropic medication purchase status and adjusting for sociodemographic and work- and health-related factors. Additionally, the statistical evidence of an overall association for total LTPA was clearly weaker than for VPA alone, which is in line with the clear null findings for LPA.

Regarding the shape of the associations, our fully adjusted results showed no statistical evidence of nonlinearity for VPA. However, the relatively small number of observations at higher VPA levels, including potentially sparse representation of some covariate values, may have limited the precision of the adjusted curve and the statistical power to detect departures from linearity. It is known that flexible spline estimates can be unstable at the tails of the exposure distribution and graphical presentations may suggest nonlinearity even though statistical significance thresholds are not met on formal tests of nonlinearity (Discacciati et al., 2025). This should also be considered in light of our substantially smaller sample size (*N* = 4,160) compared with that of Wang et al. (2025) (91,800 participants with device-measured data and 365,656 with self-reported data). Consequently, given the evidence of nonlinearity in previous studies, a nonlinear association cannot be ruled out despite the absence of statistical evidence for nonlinearity in our fully adjusted model.

Two complementary pathways may help explain our findings. First, higher-intensity physical activity may elicit greater acute physiological and neurobiological responses per unit of time than lower-intensity activity. For example, exercise intensity has been linked to acute changes in brain-derived neurotrophic factor (BDNF) and other neurobiological processes implicated in mental health (Ferris et al., 2007; Paolucci et al., 2018). More recent work views the effects of physical activity on mental health working through a range of interconnected mechanisms, including neuroplasticity, inflammation, stress-regulatory systems, neurotransmitter signaling, and endocannabinoid pathways (Budde et al., 2025; Ferris et al., 2007; Montgomery & Grant, 2026; Paolucci et al., 2018; Ross et al., 2023). It should be noted that strict causal evidence for this view or any singular neurobiological mechanism is still lacking (Budde et al., 2025). Nevertheless, the stronger associations observed for higher-intensity LTPA could partly reflect greater physiological stimulation.

A complementary interpretation is that the association between higher-intensity LTPA and better mental health may partly reflect the contexts in which activities of different intensities occur and the psychosocial experiences associated with those contexts. VPA may more often involve organized or recreational sports, such as floorball or tennis, which can provide opportunities for social interaction, enjoyment, and experiences of competence and mastery (Teixeira et al., 2012; White et al., 2024). In contrast, LPA may more often comprise routine activities embedded in daily life, such as walking to a store etc., which less frequently involve these psychosocial processes. This interpretation is consistent with emerging evidence suggesting that the mental-health effects of physical activity depend not only on how much activity people perform, but also on its intensity, modality, and social and behavioral context (Vella et al., 2023).

### Methodological considerations

Several methodological considerations should be taken into account when interpreting the results of this study. First, the observational design precludes causal inference. Second, the proportional hazards assumption was clearly violated for prior psychotropic medication purchase status, and based on visual inspection, it appeared that the association with subsequent psychotropic medication purchases weakened further into the follow-up period (Figure S2). One possible explanation is that purchases occurring early in the follow-up disproportionately represented participants with prior psychotropic medication purchases. Consequently, purchases occurring later in the follow-up may have increasingly represented participants without prior psychotropic medication use. Based on this we decided to stratify the baseline hazard by prior psychotropic medication purchase status in our main analyses.

To further investigate the impact of prior psychotropic medication status we excluded those with ongoing psychotropic medication, defined as having purchased psychotropic medication inside the 100 days leading up to the baseline survey, which attenuated the associations, although clear evidence of an association was still observed for VPA (Table S7). The weaker statistical evidence may partly reflect the smaller sample, whereas attenuation of the effect estimates may indicate confounding or selection related to ongoing psychotropic medication. However, when excluding all those with purchases in the prior 3 years, the results were still similar (Table S8). This could indicate that the association of VPA with psychotropic medication purchases is generally similar between participants with and without prior psychotropic medication purchases. Although we thoroughly examined whether prior psychotropic medication purchases influenced the observed associations, the possibility of reverse causation due to subclinical mental health problems cannot be excluded.

Third, the LTPA measures were self-reported and are therefore susceptible to recall and social desirability bias as well as reporting error. Fourth, our sample consisted predominantly of women (80.4%) and due to the small sample of men (n = 815), we conducted our main analyses in the pooled sample, with adjustment for gender.

However, our sensitivity analyses provided evidence that the LPA and VPA associations differed by gender. For VPA it seemed that the estimated hazard declined with increasing VPA among both men and women, but the decline was more pronounced for men (see Figure S4). Nevertheless, the association was inverse in both genders, which is consistent with the pooled VPA findings. Higher LPA was associated with an increased hazard among women and a decreased hazard among men. However, the gender-specific association was statistically significant only among women (Table S9). However, these differences were largely mitigated when stratifying by prior psychotropic medication purchase status and adjusting for covariates. This suggests that the associations between different intensities of LTPA and psychotropic medication purchases are differentially confounded by prior psychotropic medication and sociodemographic and work- and health-related factors between men and women. Furthermore, the study sample consists of young and early midlife employees of the City of Helsinki. Consequently, the findings should be generalized cautiously to other populations, particularly those with substantially different characteristics and contexts.

The present study also had some notable strengths. First, our outcome variable was registered psychotropic purchases based on data maintained by the Social Insurance Institution of Finland (Kela). The data from this register provide a reliable record of prescribed, reimbursed psychotropic medication purchases in the study population. Furthermore, the register provided data until the end of 2022 allowing up to approximately five years of follow-up. Additionally, we had a large sample that allowed the examination of nonlinear associations between different intensities of LTPA and subsequent psychotropic medication purchases while adjusting for multiple potential confounders. Lastly, multiple sensitivity analyses were performed which generally supported the robustness of the findings.

## Conclusion

The present study provides evidence that associations between LTPA and subsequent psychotropic medication purchases differ by activity intensity. Higher-intensity activity, particularly VPA, was more clearly associated with a lower hazard of a first psychotropic medication purchase. In absolute terms, 5 hours/week of VPA was associated with approximately 23 to 78, depending on the level of adjustment, fewer participants experiencing a first psychotropic medication purchase per 1,000 over three years compared with no VPA. The results gave some indication of diminishing returns, with the estimated association for VPA being steepest at lower activity levels and appearing to level off at approximately 5 hours/week. However, evidence of diminishing returns was not retained after full adjustment. A diminishing-returns pattern was also observed for MPA, although the overall association did not receive clear statistical support. The promotion of leisure-time MVPA, preferably at a vigorous intensity, may be an important target for reducing the burden caused by mental health issues as indicated by psychotropic medication purchases. Future studies using device-measured physical activity and larger and more gender-balanced samples are needed to clarify the dose–response shapes and potential differences between activity intensities.

## AI statement

Generative artificial intelligence was used to check and improve the language of the manuscript and to edit selected supplementary plot axis labels. All AI-assisted changes were reviewed and approved by the authors, who take full responsibility for the final content.

## CRediT authorship contribution statement

**Lari Järvensivu:** Conceptualization, Data curation, Formal analysis, Methodology, Validation, Visualization, Writing – original draft, Writing – review & editing. **Ville Päivärinne:** Conceptualization, Supervision, Writing – review & editing. **Ossi Rahkonen:** Conceptualization, Funding acquisition, Supervision, Writing – review & editing. **Tea Lallukka:** Conceptualization, Supervision, Writing – review & editing.

## Ethics approval and consent to participate

The Helsinki Health Study protocol was reviewed by the Ethics Committee of the Faculty of Medicine, University of Helsinki, which issued a favourable statement in February 2017 (Decision No. 01/2017). The City of Helsinki granted additional approval in April 2017 (Decision No. HEL 2017-004490), and the City of Helsinki health and personnel authorities issued the research permit (TK/470/07.03.00/2023). This study was restricted to participants who had provided informed consent for register linkage and was conducted in Kapseli®, a secure data-processing environment provided by the Finnish Social and Health Data Permit Authority (Findata). The study was conducted in accordance with applicable ethical guidelines and data-protection requirements.

## Data availability statement

Due to data protection requirements and the sensitive nature of the data, the data cannot be made publicly available. The data protection statement is available at https://www.helsinki.fi/en/researchgroups/helsinki-health-study/data-protection-statement.

