## Supplemental file for "Associations between different intensities of leisure-time physical activity and subsequent psychotropic medication use among young and early midlife adults: a register-linked follow-up study"

Table S1.
*Metabolic equivalent multipliers used to calculate MET-hours for different intensities of leisure-time physical activity.*

| **Activity intensity** | **Intensity label** | **MET multiplier** | **MET-hours calculation** |
|---|---|---|---|
| Walking or other equal-intensity activities | Light | 2.8 | Hours × 2.8 |
| Brisk walking or other equal-intensity activities | Moderate | 4.3 | Hours × 4.3 |
| Jogging or other equal-intensity activities | Low-vigorous | 7.0 | Hours × 7.0 |
| Running or other equal-intensity activities | High-vigorous | 9.0 | Hours × 9.0 |

*Note.* LTPA = leisure-time physical activity; MET = metabolic equivalent; MPA = moderate physical activity; MVPA = moderate-to-vigorous physical activity; VPA = vigorous physical activity. Total LTPA was calculated by combining light, moderate, low-vigorous, and high-vigorous physical activity. VPA was calculated by combining low-vigorous and high-vigorous physical activity. MVPA was calculated by combining MPA and VPA.

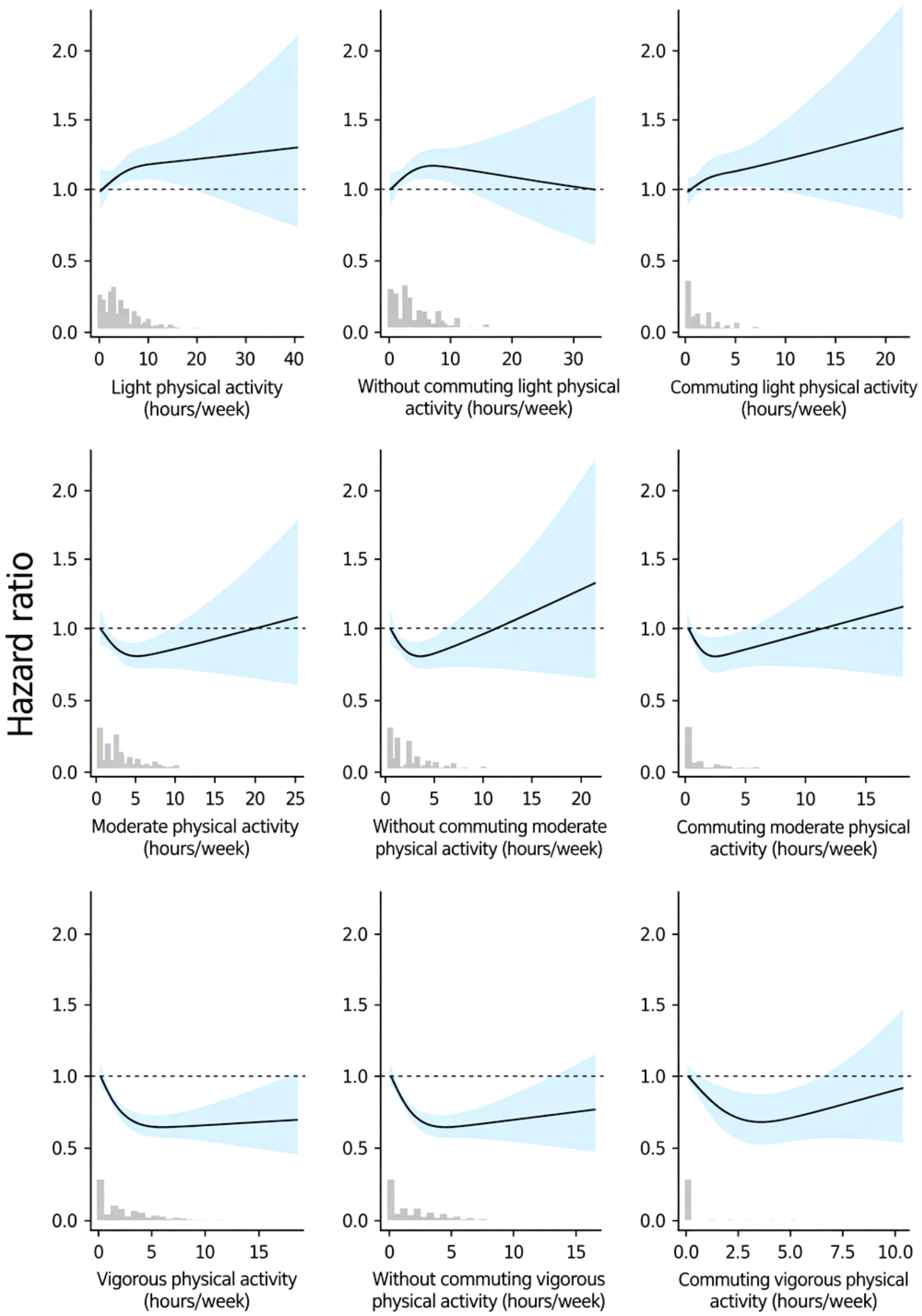


Figure S1. *Associations of leisure-time physical activity with the hazard of a first psychotropic medication purchase, separately for total leisure-time physical activity, leisure-time physical activity excluding commuting, and commuting physical activity only, among 19–39-year-old participants of the Helsinki Health Study (N = 4,160).*

*Note.* LPA = light physical activity; LTPA = leisure-time physical activity; MPA = moderate physical activity; VPA = vigorous physical activity. From left to right, the panels present total LTPA, LTPA excluding commuting, and commuting physical activity only. Estimates are from Model 1, which was adjusted for age and gender.

Table S2.

*Pearson correlations among light, moderate, and vigorous leisure-time physical activity among 19–39-year-old participants of the Helsinki Health Study (N = 4,160)*

| **Variable** | **1** | **2** | **3** |
|---|---|---|---|
| 1. Light physical activity (hours/week) | — | | |
| 2. Moderate physical activity (hours/week) | .40 | — | |
| 3. Vigorous physical activity (hours/week) | .11 | .16 | — |

Table S3.

*Generalized variance inflation factors for different intensities of leisure-time physical activity and covariates among 19–39-year-old participants of the Helsinki Health Study (N = 4,160)*

| Predictor | df | GVIF | Adjusted GVIF ($\mathrm{GVIF}^{1/(2\times\mathrm{df})}$) |
|---|---|---|---|
| Light physical activity | 2 | 18.09 | 2.06 |
| Moderate physical activity | 2 | 16.52 | 2.02 |
| Vigorous physical activity | 2 | 14.93 | 1.97 |
| Prior psychotropic purchase: Yes | 1 | 1.05 | 1.02 |
| Gender: woman | 1 | 1.16 | 1.08 |
| Age | 1 | 1.16 | 1.08 |
| Education: Intermediate | 1 | 1.45 | 1.20 |
| Education: High | 1 | 1.73 | 1.32 |
| Marital status | 1 | 1.06 | 1.03 |
| Smoking | 1 | 1.14 | 1.07 |
| Binge drinking | 1 | 1.09 | 1.04 |
| Fresh vegetables | 1 | 1.12 | 1.06 |
| Sedentary time | 1 | 1.09 | 1.04 |
| Body mass index | 1 | 1.13 | 1.06 |
| Physical strenuousness of work: Low | 1 | 1.32 | 1.15 |
| Mental strenuousness of work: Low | 1 | 1.05 | 1.03 |

*Note. df = degrees of freedom; GVIF = generalized variance inflation factor;* $\mathrm{GVIF}^{1/(2\times\mathrm{df})}$*= degrees-of-freedom-adjusted generalized variance inflation factor.*

Table S4.

*Grambsch–Therneau tests of the proportional hazards assumption for the fully adjusted Cox proportional hazards model among 19–39-year-old participants of the Helsinki Health Study (N = 4,160)*

| Model term | $\chi^2$ | df | *p* |
|---|---|---|---|
| Light physical activity | 7.25 | 2 | 0.0267 |
| Moderate physical activity | 0.91 | 2 | 0.6361 |
| Vigorous physical activity | 0.28 | 2 | 0.8682 |
| Prior psychotropic medication purchases | 156.62 | 1 | <0.0001 |
| Gender | 5.17 | 1 | 0.023 |
| Age | 6.99 | 1 | 0.0082 |
| Education | 0.39 | 2 | 0.8225 |
| Marital status | 5.47 | 1 | 0.0193 |
| Smoking | 10.63 | 1 | 0.0011 |
| Binge drinking | 0.08 | 1 | 0.7709 |
| Fresh vegetable intake | 3.64 | 1 | 0.0565 |
| Sedentary time | 1.11 | 1 | 0.292 |
| Body mass index | 3.05 | 1 | 0.0805 |
| Physical strenuousness of work | 9.95 | 1 | 0.0016 |
| Mental strenuousness of work | 0.67 | 1 | 0.4141 |

*Note.* $\chi^2$ = chi-square statistic; df = degrees of freedom; p = p-value. The Grambsch–Therneau test assesses whether the scaled Schoenfeld residuals are associated with follow-up time. A statistically significant p value indicates evidence against the proportional hazards assumption.

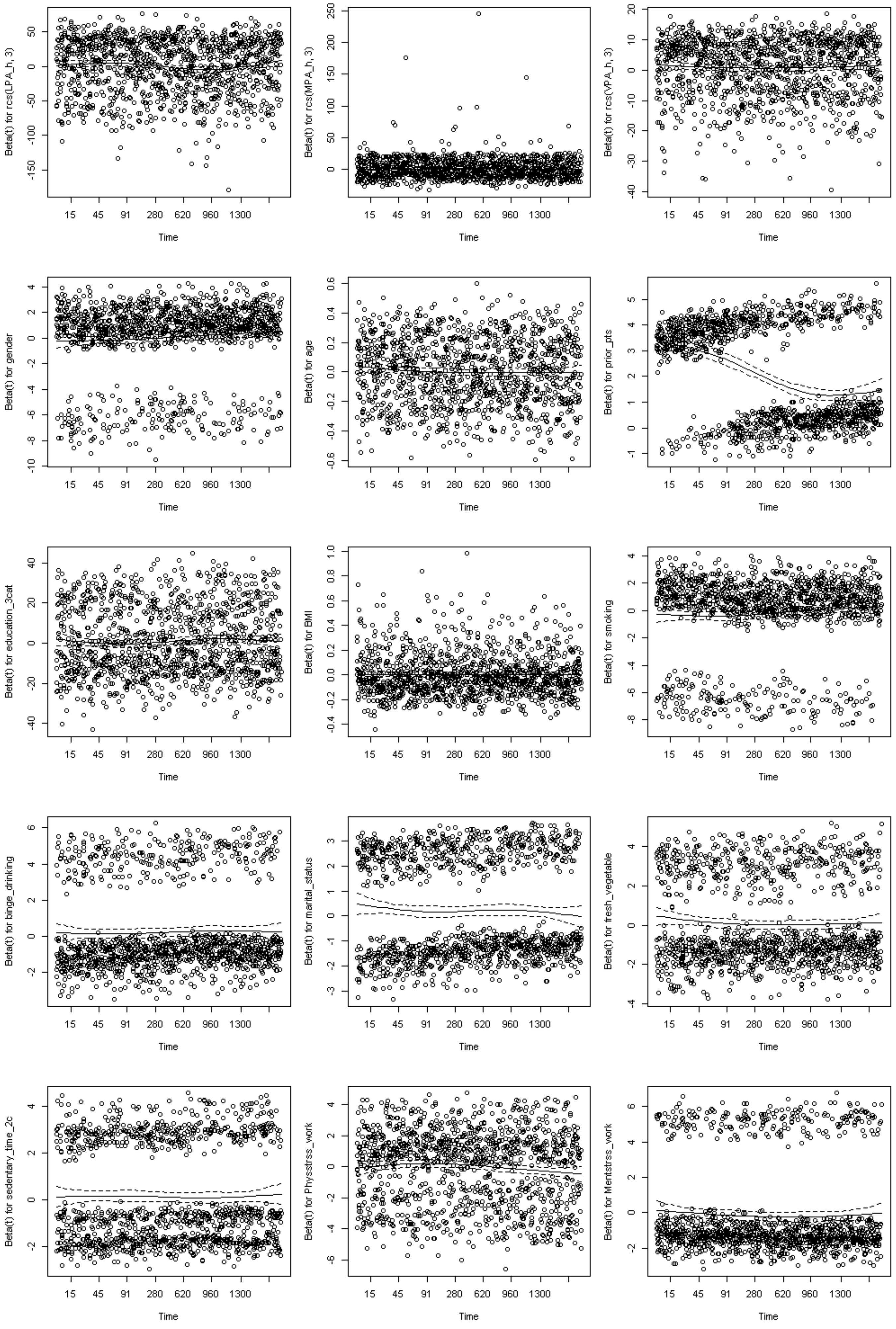
Beta(t) for rcs(LPA_h, 3)
Beta(t) for rcs(MPA_h, 3)
Beta(t) for rcs(VPA_h, 3)
Beta(t) for gender
Beta(t) for age
Beta(t) for prior_pts
Beta(t) for education_3cat
Beta(t) for BMI
Beta(t) for smoking
Beta(t) for binge_drinking
Beta(t) for marital_status
Beta(t) for fresh_vegetable
Beta(t) for sedentary_time_2c
Beta(t) for Physstrss_work
Beta(t) for Mentstrss_work
Time
15 45 91 280 620 960 1300

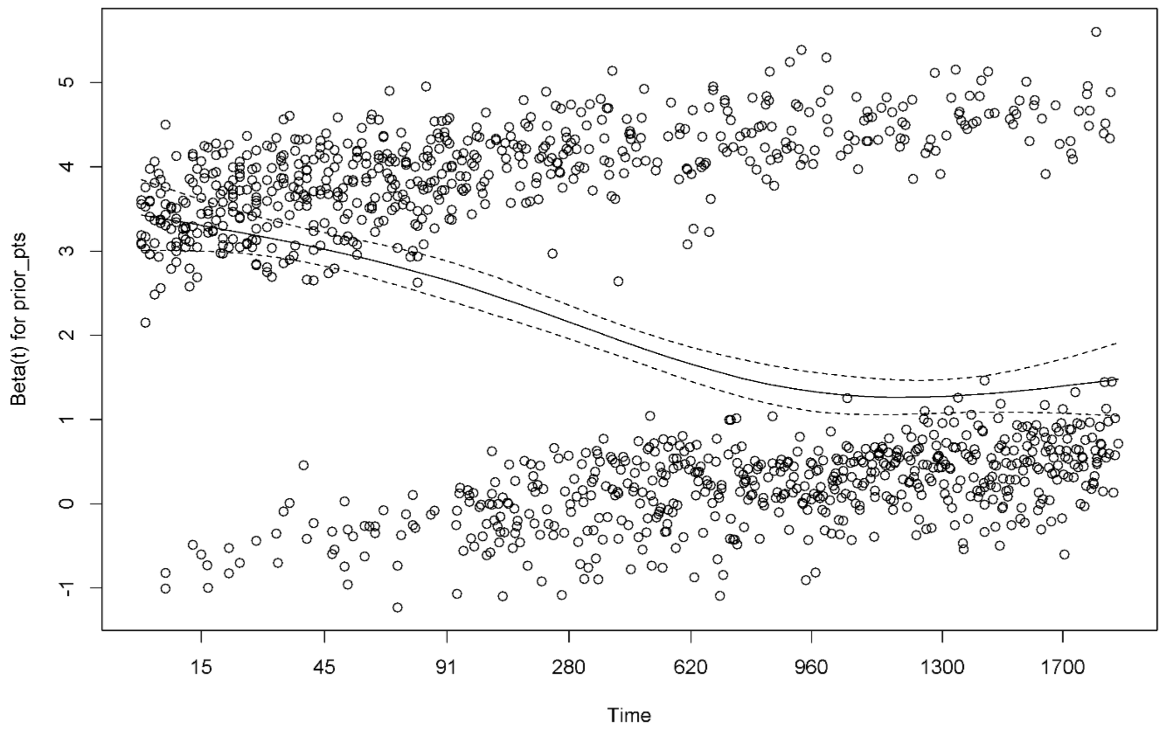


Figure S2. *Scaled Schoenfeld residual plots for the fully adjusted Cox proportional hazards model among 19–39-year-old participants of the Helsinki Health Study (N = 4,160)*

*Note*. BMI = body mass index; LPA = light physical activity; MPA = moderate physical activity; VPA = vigorous physical activity. The panels show scaled Schoenfeld residuals for each model term plotted against follow-up time. The solid line represents the smoothed relationship between the scaled Schoenfeld residuals and follow-up time. The shaded area represents the 95% confidence intervals. A clear systematic change in the residuals over follow-up time may indicate a violation of the Cox proportional hazards assumption.

Table S5.

*Overall and nonlinear Wald tests for restricted cubic spline terms in Cox proportional hazards models after excluding values at or above the 99th percentile of light, moderate, or vigorous leisure-time physical activity among 19–39-year-old participants of the Helsinki Health Study (N = 4,054)*

| Exposure | Overall Wald $\chi^2$ | df | *p* | Nonlinear Wald $\chi^2$ | df | *P* |
|---|---|---|---|---|---|---|
| **Model 1** | | | | | | |
| LPA (hours/week) | 2.92 | 2 | 0.2321 | 1.91 | 1 | 0.1665 |
| MPA (hours/week) | 4.36 | 2 | 0.1133 | 1.96 | 1 | 0.1619 |
| VPA (hours/week) | 32.61 | 2 | <0.0001 | 13.24 | 1 | 0.0003 |
| **Model 2** | | | | | | |
| LPA (hours/week) | 1.10 | 2 | 0.5764 | 1.04 | 1 | 0.3072 |
| MPA (hours/week) | 2.82 | 2 | 0.2446 | 2.33 | 1 | 0.1272 |
| VPA (hours/week) | 16.68 | 2 | 0.0002 | 5.63 | 1 | 0.0177 |
| **Model 3** | | | | | | |
| LPA (hours/week) | 1.13 | 2 | 0.5685 | 1.08 | 1 | 0.2982 |
| MPA (hours/week) | 1.11 | 2 | 0.5733 | 0.93 | 1 | 0.3359 |
| VPA (hours/week) | 6.80 | 2 | 0.0335 | 2.97 | 1 | 0.0847 |

*Note. $\chi^2$ = Wald chi-square statistic; p = p-value; df= degrees of freedom; LPA = light physical activity; MPA = moderate physical activity; VPA = vigorous physical activity. Overall p-values test the overall association of each restricted cubic spline term with the outcome. Nonlinear p values test the nonlinear spline components. Model 1 was adjusted for age and gender. Model 2 was adjusted for age and gender, with the baseline hazard stratified by prior psychotropic medication purchase status. Model 3 was adjusted for age, gender, education, body mass index, smoking, binge drinking, marital status, fresh vegetable intake, sedentary time, physical strenuousness of work, and mental strenuousness of work, with the baseline hazard stratified by prior psychotropic medication purchase status.*

Table S6.

*Overall and nonlinear Wald tests for restricted cubic spline terms in Cox proportional hazards models for moderate-to-vigorous and total leisure-time physical activity among*

| Exposure | Overall Wald $\chi^2$ | df | p | Nonlinear Wald $\chi^2$ | df | p |
|---|---|---|---|---|---|---|
| **Model 1a** | | | | | | |
| LPA (MET-hours/week) | 4.43 | 2 | 0.1094 | 0.70 | 1 | 0.4028 |
| MVPA (MET-hours/week) | 39.06 | 2 | <0.0001 | 22.05 | 1 | <0.0001 |
| **Model 1b** | | | | | | |
| Total LTPA (MET-hours/week) | 21.17 | 2 | <0.0001 | 14.95 | 1 | 0.0001 |
| **Model 2a** | | | | | | |
| LPA (MET-hours/week) | 1.79 | 2 | 0.4093 | 0.22 | 1 | 0.6412 |
| MVPA (MET-hours/week) | 20.34 | 2 | <0.0001 | 7.21 | 1 | 0.0073 |
| **Model 2b** | | | | | | |
| Total LTPA (MET-hours/week) | 10.70 | 2 | 0.0047 | 3.85 | 1 | 0.0496 |
| **Model 3a** | | | | | | |
| LPA (MET-hours/week) | 0.56 | 2 | 0.7571 | 0.24 | 1 | 0.6271 |
| MVPA (MET-hours/week) | 8.41 | 2 | 0.0149 | 2.31 | 1 | 0.1284 |
| **Model 3b** | | | | | | |
| Total LTPA (MET-hours/week) | 4.74 | 2 | 0.0937 | 0.60 | 1 | 0.4373 |

*Note*. $\chi^2$ = Wald chi-square statistic; df = degrees of freedom; p = p-value; MET = metabolic equivalent LPA = light physical activity; LTPA = leisure-time physical activity; MVPA = moderate-to-vigorous physical activity. Overall p values test the overall association of each restricted cubic spline term with the outcome. Nonlinear p values test the nonlinear spline components. Model 1 was adjusted for age and gender. Model 2 was adjusted for age and gender, with the baseline hazard stratified by prior psychotropic medication purchase status. Model 3 was adjusted for age, gender, education, body mass index, smoking, binge drinking, marital status, fresh vegetable intake, sedentary time, physical strenuousness of work, and mental strenuousness of work, with the baseline hazard stratified by prior psychotropic medication purchase status. In the MVPA models, LPA and MVPA were mutually adjusted. Total LTPA was modelled as a single exposure.

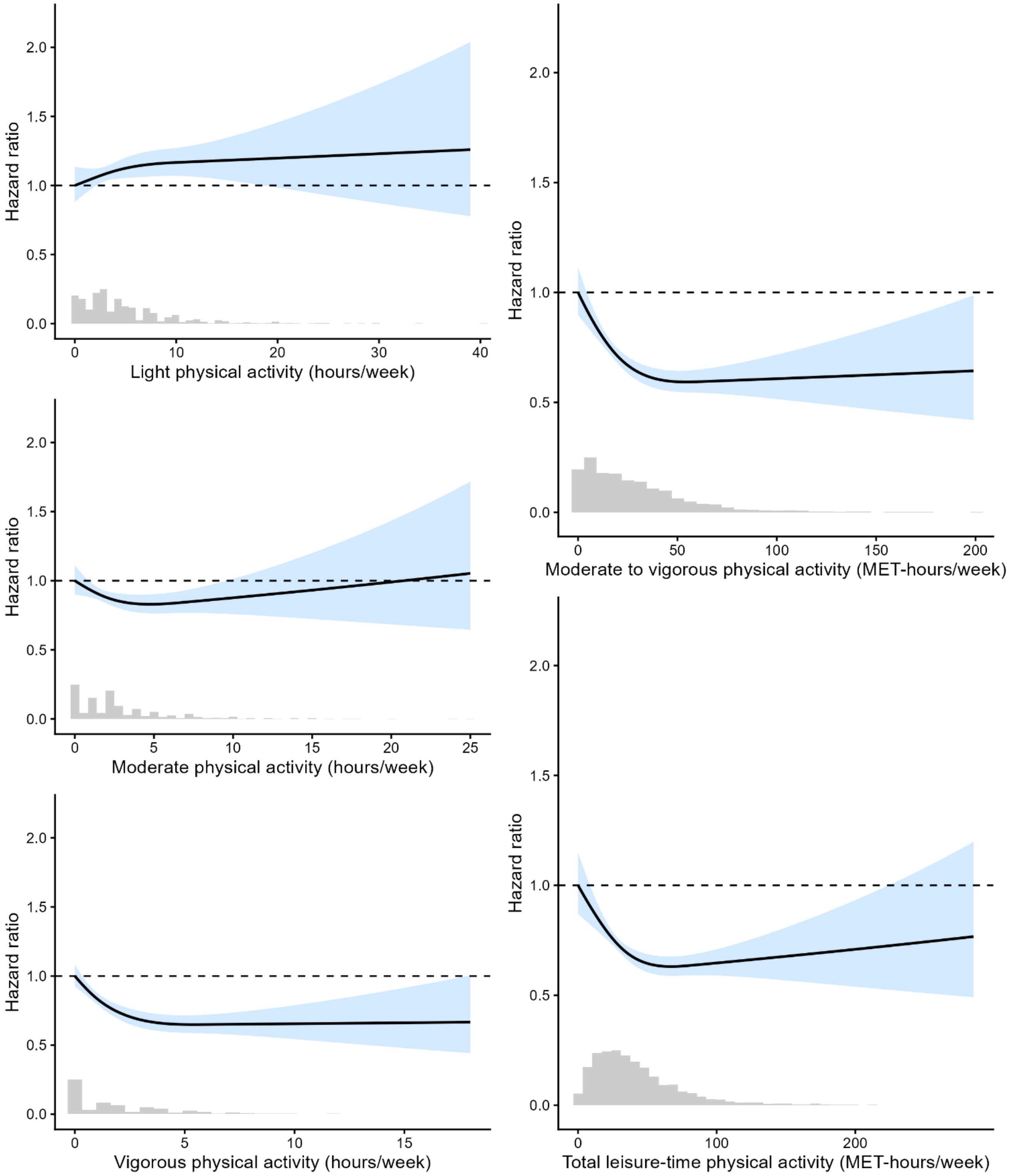


Figure S3. *Model 1 associations of light, moderate, vigorous, moderate-to-vigorous, and total leisure-time physical activity with the hazard of a first psychotropic medication purchase among 19–39-year-old participants of the Helsinki Health Study (N = 4,160)*

*Note*. Blue bands indicate 95% confidence intervals for the hazard ratios. Histograms depict the distribution of participants across values of hours per week or MET-hours per week, as applicable. Model 1 was adjusted for age and gender. LPA = light physical activity;

LTPA = leisure-time physical activity; MET = metabolic equivalent; MPA = moderate physical activity; MVPA = moderate-to-vigorous physical activity; VPA = vigorous physical activity.

Table S7.

*Overall and nonlinear Wald tests for restricted cubic spline terms in Cox proportional hazards models among 19–39-year-old participants of the Helsinki Health Study without ongoing psychotropic medication use (N = 3,826)*

| Exposure | Overall Wald $\chi^2$ | df | *p* | Nonlinear Wald $\chi^2$ | df | *p* |
|---|---|---|---|---|---|---|
| **Model 1** | | | | | | |
| LPA (hours/week) | 2.97 | 2 | 0.2261 | 0.02 | 1 | 0.8923 |
| MPA (hours/week) | 1.73 | 2 | 0.4203 | 1.54 | 1 | 0.2142 |
| VPA (hours/week) | 15.46 | 2 | 0.0004 | 5.58 | 1 | 0.0181 |
| **Model 2** | | | | | | |
| LPA (hours/week) | 1.35 | 2 | 0.5083 | 0.00 | 1 | 0.9592 |
| MPA (hours/week) | 1.37 | 2 | 0.5032 | 1.35 | 1 | 0.2447 |
| VPA (hours/week) | 13.02 | 2 | 0.0015 | 1.86 | 1 | 0.1721 |
| **Model 3** | | | | | | |
| LPA (hours/week) | 0.65 | 2 | 0.7221 | 0.00 | 1 | 0.9566 |
| MPA (hours/week) | 0.50 | 2 | 0.7769 | 0.50 | 1 | 0.4794 |
| VPA (hours/week) | 6.77 | 2 | 0.0340 | 0.50 | 1 | 0.4803 |

*Note*. $\chi^2$ = Wald chi-square statistic; df = degrees of freedom; p = p-value; LPA = light physical activity; MPA = moderate physical activity; VPA = vigorous physical activity. Overall p-values test the overall association of each restricted cubic spline term with the outcome. Nonlinear p-values test the nonlinear spline components. Ongoing psychotropic medication use was defined as a psychotropic medication purchase during the 100 days before the survey response. Model 1 was adjusted for age and gender. Model 2 was adjusted for age and gender, with the baseline hazard stratified by prior psychotropic medication purchase status. Model 3 was adjusted for age, gender, education, body mass index, smoking, binge drinking, marital status, fresh vegetable intake, sedentary time, physical strenuousness of work, and mental strenuousness of work, with the baseline hazard stratified by prior psychotropic medication purchase status.

Table S8.

*Overall and nonlinear Wald tests for restricted cubic spline terms in Cox proportional hazards models among 19–39-year-old participants of the Helsinki Health Study without psychotropic medication purchases during the three years before the 2017 survey (N = 3,371)*

| Exposure | Overall Wald $\chi^2$ | df | *p* | Nonlinear Wald $\chi^2$ | df | *p* |
|---|---|---|---|---|---|---|
| **Model 1** | | | | | | |
| LPA (hours/week) | 1.57 | 2 | 0.457 | 0.04 | 1 | 0.841 |
| MPA (hours/week) | 1.03 | 2 | 0.599 | 0.32 | 1 | 0.571 |
| VPA (hours/week) | 13.96 | 2 | 0.001 | 6.14 | 1 | 0.013 |
| **Model 2** | | | | | | |
| LPA (hours/week) | 1.36 | 2 | 0.507 | 0.02 | 1 | 0.875 |
| MPA (hours/week) | 1.14 | 2 | 0.566 | 0.07 | 1 | 0.787 |
| VPA (hours/week) | 7.22 | 2 | 0.027 | 3.05 | 1 | 0.081 |

*Note*. $\chi^2$ = Wald chi-square statistic; df = degrees of freedom; p = p-value; LPA = light physical activity; MPA = moderate physical activity; VPA = vigorous physical activity. Overall p values test the overall association of each restricted cubic spline term with the outcome. Nonlinear p values test the nonlinear spline components. Model 1 was adjusted for age and gender. Model 2 was adjusted for age, gender, education, body mass index, smoking, binge drinking, marital status, fresh vegetable intake, sedentary time, physical strenuousness of work, and mental strenuousness of work.

Table S9.

*Interactions of light, moderate, and vigorous leisure-time physical activity with gender and gender-specific associations with the hazard of a first psychotropic medication purchase among 19–39-year-old participants of the Helsinki Health Study (N = 4,160; men, n = 815; women, n = 3,345)*

| Exposure | Overall Interaction Wald χ² | df | p-value | Nonlinear Interaction Wald χ² | df | p-value |
|---|---|---|---|---|---|---|
| **Model 1** | | | | | | |
| LPA (hours/week) × gender | 6.82 | 2 | 0.0330 | 4.94 | 1 | 0.0262 |
| MPA (hours/week) × gender | 3.91 | 2 | 0.1417 | 3.51 | 1 | 0.0610 |
| VPA (hours/week) × gender | 6.50 | 2 | 0.0387 | 3.27 | 1 | 0.0706 |
| **Model 2** | | | | | | |
| LPA (hours/week) × gender | 0.94 | 2 | 0.6236 | 0.26 | 1 | 0.6103 |
| MPA (hours/week) × gender | 1.53 | 2 | 0.4662 | 0.41 | 1 | 0.5244 |
| VPA (hours/week) × gender | 3.61 | 2 | 0.1642 | 1.16 | 1 | 0.2819 |
| **Model 3** | | | | | | |
| LPA (hours/week) × gender | 0.45 | 2 | 0.7967 | 0.07 | 1 | 0.7929 |
| MPA (hours/week) × gender | 1.33 | 2 | 0.5133 | 0.45 | 1 | 0.5018 |
| VPA (hours/week) × gender | 2.88 | 2 | 0.237 | 1.05 | 1 | 0.3058 |

| | **Men**: overall Wald χ² | df | p-value | **Women**: overall Wald χ² | df | p-value |
|---|---|---|---|---|---|---|
| **Model 1** | | | | | | |
| LPA (hours/week) | 3.16 | 2 | 0.2064 | 6.27 | 2 | 0.0434 |
| MPA (hours/week) | 0.84 | 2 | 0.6561 | 8.03 | 2 | 0.0180 |
| VPA (hours/week) | 23.17 | 2 | <0.0001 | 18.37 | 2 | 0.0001 |
| **Model 2** | | | | | | |
| LPA (hours/week) | 0.34 | 2 | 0.8442 | 1.4 | 2 | 0.4961 |
| MPA (hours/week) | 1.04 | 2 | 0.5953 | 4.59 | 2 | 0.1007 |
| VPA (hours/week) | 14.77 | 2 | 0.0006 | 11.65 | 2 | 0.0029 |
| **Model 3** | | | | | | |
| LPA (hours/week) | 0.32 | 2 | 0.8516 | 0.61 | 2 | 0.7366 |
| MPA (hours/week) | 0.9 | 2 | 0.638 | 2.58 | 2 | 0.2749 |
| VPA (hours/week) | 9.46 | 2 | 0.0088 | 5.14 | 2 | 0.0764 |

*Note*. χ² = Wald chi-square statistic; df = degrees of freedom; p = p-value; LPA = light physical activity; MPA = moderate physical activity; VPA = vigorous physical activity. Overall p values test the overall association of each restricted cubic spline term with the outcome. Overall interaction p values test whether the overall association differs between men and women. Nonlinear interaction p values test whether the nonlinear component of the association differs between men and women. Model 1 was adjusted for age and gender. Model 2 was adjusted for age and gender, with the baseline hazard stratified by prior psychotropic medication purchase status. Model 3 was adjusted for age, gender, education, body mass index, smoking, binge drinking, marital status, fresh vegetable intake, sedentary time, physical strenuousness of work, and mental strenuousness of work, with the baseline hazard stratified by prior psychotropic medication purchase status.

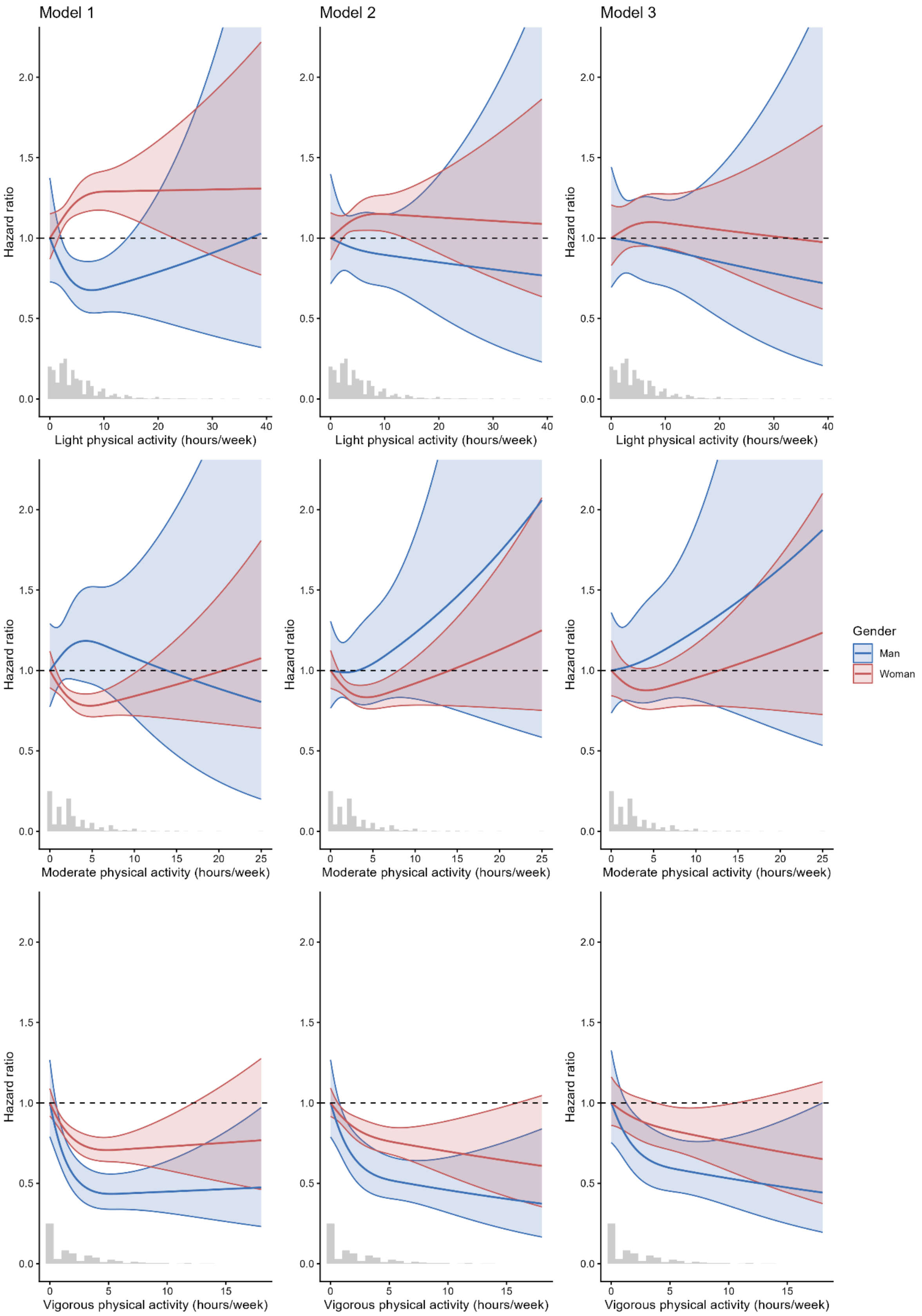


Figure S4. *Gender-specific associations of light, moderate, and vigorous leisure-time physical activity with the hazard of a first psychotropic medication purchase among 19–39-year-old participants of the Helsinki Health Study (N = 4,160; men, n = 815; women, n = 3,345)*

*Note*. Blue and red bands indicate 95% confidence intervals for the hazard ratios for men and women, respectively. Histograms depict the distribution of participants across values of light, moderate, and vigorous physical activity. Estimates were obtained from gender interaction models. Model 1 was adjusted for age and gender. Model 2 was adjusted for age and gender, with the baseline hazard stratified by prior psychotropic medication purchase status. Model 3 was adjusted for age, gender, education, body mass index, smoking, binge drinking, marital status, fresh vegetable intake, sedentary time, physical strenuousness of work, and mental strenuousness of work, with the baseline hazard stratified by prior psychotropic medication purchase status.

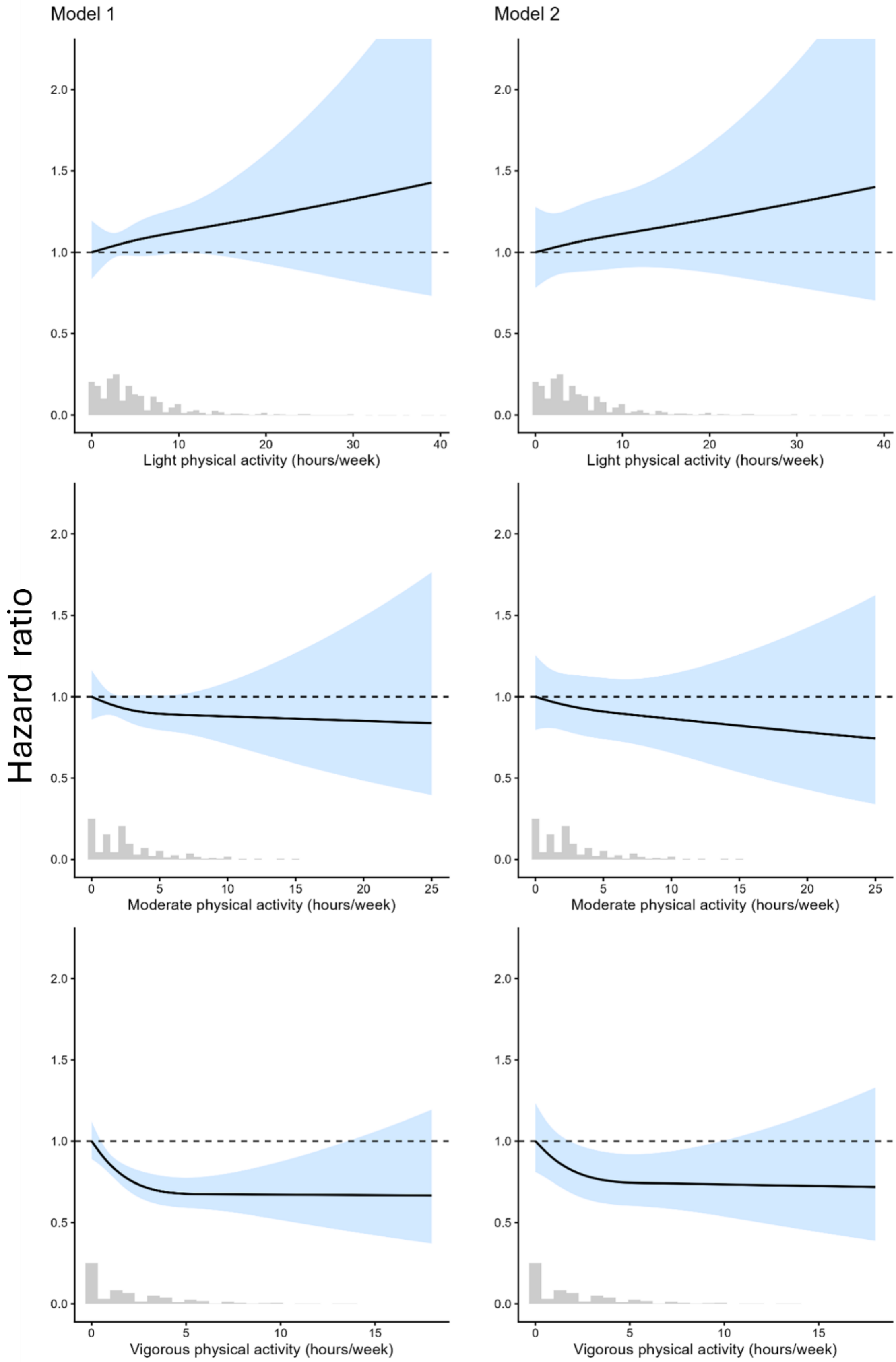
Model 1
Model 2
Hazard ratio
Light physical activity (hours/week)
Moderate physical activity (hours/week)
Vigorous physical activity (hours/week)
2.0
1.5
1.0
0.5
0.0
0
10
20
30
40
5
15
25

Figure S5. *Associations of light, moderate, and vigorous leisure-time physical activity with the hazard of a first psychotropic medication purchase among 19–39-year-old participants of the Helsinki Health Study without psychotropic medication purchases during the three years before the 2017 survey (N = 3,371)*

*Note*. Blue bands indicate 95% confidence intervals for the hazard ratios. Histograms depict the distribution of participants across values of light, moderate, and vigorous physical activity. A total of 789 participants with psychotropic medication purchases during the three years before the 2017 survey were excluded. Model 1 was adjusted for age and gender. Model 2 was adjusted for age, gender, education, body mass index, smoking, binge drinking, marital status, fresh vegetable intake, sedentary time, physical strenuousness of work, and mental strenuousness of work.
